# Florzolotau (18F) PET provides in-vivo measures of tau pathology in progressive supranuclear palsy: An imaging-postmortem correlation study

**DOI:** 10.64898/2025.12.02.25340863

**Authors:** Nobuyuki Araki, Takahiro Takeda, Yuhei Takado, Kenji Tagai, Erika Seki, Nobutaka Arai, Sagiri Isose, Kimiko Ito, Kimihito Arai, Kazuhiro Honda, Satoshi Kuwabara, Makoto Higuchi, Hironobu Endo

## Abstract

The clinical diagnosis of 4-repeat tauopathies, including progressive supranuclear palsy (PSP) and corticobasal degeneration, remains challenging. Although patients may present with characteristic clinical features suggestive of these diseases, discrepancies frequently arise between antemortem diagnosis and postmortem pathological findings. Reliable biomarkers for in vivo detection of tau deposition are therefore urgently needed. We report a case of a man in his 60s who developed progressive gait disturbance and cerebellar ataxia. Based on clinical findings, PSP with predominant cerebellar ataxia (PSP-C) was suspected, and tau positron emission tomography (PET) using florzolotau (18F) ([^18^F]florzolotau) was performed.

The PET scan revealed marked tracer retention in the upper brainstem, bilateral diencephalon, and basal ganglia, consistent with the PSP-C phenotype. The patient died of aspiration pneumonia three years after the first visit, and a neuropathological examination confirmed PSP, showing neuronal and glial tau aggregates, including tufted astrocytes, primarily in the midbrain, pons, dentate nucleus and subthalamic nucleus.

To quantitatively assess the concordance between in vivo and postmortem findings, tau burden was measured across 48 representative brain regions using AT8 immunohistochemistry and a binarization-based quantification method. Regional tau pathology presented a strong positive correlation with the standardized uptake value ratio of [^18^F]florzolotau derived from the antemortem PET data.

In conclusion, this study provides direct evidence that [^18^F]florzolotau-PET accurately reflects the regional distribution of tau pathology in PSP. Our findings support the clinical utility of [^18^F]florzolotau as a sensitive probe for diagnosing 4-repeat tauopathies and for therapeutic evaluations targeting these disorders.

## Introduction

The radiotracer florzolotau (18F) ([^18^F]florzolotau), also known as [^18^F]PM-PBB3 and [^18^F]APN-1607, enables high-contrast positron emission tomography (PET) imaging of fibrillar tau pathologies in living humans. This radioligand is capable of visualizing not only Alzheimer’s disease (AD)-type tau aggregates but also 4-repeat (4R) tau pathologies, as seen in progressive supranuclear palsy (PSP) and corticobasal degeneration (CBD), and 3-repeat (3R) tau deposits characteristic of Pick’s disease [1]. The regional distribution and severity of tau pathology within the central nervous system closely correlate with disease progression and symptomatology [2]. Consistently, the extent and intensity of [^18^F]florzolotau uptake have been shown to associate with clinical severity of AD and PSP [1].

While [^18^F]florzolotau-PET holds great promise for improving diagnostic accuracy in 4R tauopathies, the degree to which in vivo tracer retention reflects underlying neuropathological tau burden remains incompletely understood [3, 4]. Quantitative imaging-neuropathological correlations have only been documented with this tracer in a case with Pick’s disease [5], despite demonstrating its ability to capture disease-specific tau topologies in autopsy-confirmed PSP and biopsy-proven CBD [1]. To clarify the pathological relevance of [^18^F]florzolotau retention, it is essential to perform parallel in vivo PET imaging and postmortem histopathological analyses in the same individuals.

PSP comprises several clinical subtypes, some of which are difficult to distinguish from other movement disorders. Among these, PSP with predominant cerebellar ataxia (PSP-C) represents a rare and diagnostically challenging phenotype [6]. Here, we describe a patient with PSP-C who underwent [^18^F]florzolotau-PET nearly two years before death. By quantitatively comparing in vivo PET signals with postmortem pathology, we aimed to evaluate the capability of this tau PET technique to accurately reflect regional tau deposition. This case provides direct evidence supporting the clinical utility of [^18^F]florzolotau-PET for estimating tau burden in living cases with 4R tauopathies.

### Case presentation

A previously healthy man in his 60s developed gait disturbance with frequent backward falls. His gait initially appeared shuffling but gradually evolved into a calcaneal pattern. Around the same time, he began to exhibit memory lapses, occasional irritability toward his wife over trivial matters, and dysarthria. His facial expression became mask-like, and his eyes appeared vacant. He was referred to the Neurology Department of the National Hospital Organization Chiba Medical Center Chibahigashi National Hospital at the same age as he had been at his first visit. Classical parkinsonian symptoms such as resting tremors and cogwheel rigidity were absent.

His dysarthria progressively worsened, and he developed clumsiness in using chopsticks and increasingly illegible handwriting. Given the combination of severe ocular motor impairment and gait disturbance, PSP was suspected. there was no family history of neurological disorders, although his parents were first cousins. Ocular examination revealed central fixation with limited bilateral adduction, while doll’s eye phenomenon was preserved. Speech was slurred and irregular in rhythm. No rigidity was observed in the bilateral upper limbs or trunk, but mild spasticity was present in both lower limbs, and tendon reflexes were exaggerated in all extremities. Left-dominant mild dysmetria and motor decomposition were noted during finger-nose and heel-knee tests, whereas sensory function remained intact. He could stand without assistance, and Romberg’s sign was negative; however, his gait showed lateral sway with posterior displacement of the center of gravity. Arm swing was increased, and his gait stance was wide. Myerson’s sign was positive, but the applause sign was absent. The Scale for the Assessment and Rating of Ataxia was 9.5/40, and the Unified Parkinson’s Disease Rating Scale-Part 3 score was 7/108.

Routine blood and biochemical tests, including thyroid function, were within normal limits. No autoantibodies indicative of autoimmune diseases were detected. β-hexosaminidase activity, very-long-chain fatty acids, and oxysterol levels were normal. Cerebrospinal fluid analysis, including tau and amyloid-β (Aβ) quantification, yielded normal results. Genetic testing for spinocerebellar ataxia was negative. Brain magnetic resonance imaging (MRI) showed atrophy of the tegmentum mesencephali, with mild cerebellar and pontine tegmentum atrophy (Figure 1). N-isopropyl-p-[^123^I]iodoamphetamine single-photon emission computed tomography (SPECT) demonstrated reduced perfusion in the bilateral frontal cortex, with preserved flow in the basal ganglia and mild reduction in the cerebellum. Dopamine transporter SPECT revealed reduced uptake in the bilateral striata (right specific binding ratio [SBR], 2.07; left SBR, 1.81). [^123^I]-meta-iodobenzylguanidine cardiac scintigraphy showed a normal heart-to-mediastinum ratio (early, 3.13; delayed, 3.31). One year after the first visit, tau PET using [^18^F]florzolotau and amyloid PET using [^11^C]Pittsburgh Compound-B were performed (see neuroradiology section).

**Figure 1.**
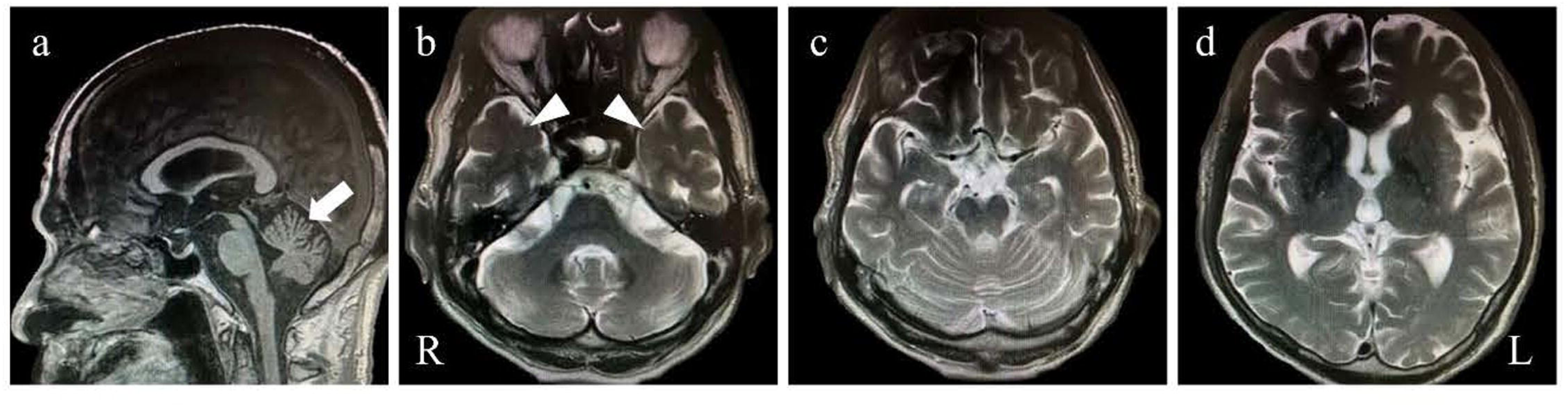
MRI findings. Sagittal T1WIs show marked atrophy of the midbrain tegmentum (a) and mild atrophy of the cerebellar dorsal vermis (arrow, b). The medial temporal lobes (arrowheads) are relatively preserved (c). The basal ganglia appear normal (c), while the cerebral cortex exhibits mild atrophy (d). L, left; R, right.

Based on these findings, the diagnosis of PSP-C was made [6, 7]. Over time, his falls became more frequent, and muscle rigidity emerged, fulfilling the Movement Disorder Society new diagnostic criteria for PSP-Richardson [8]. Two years after the first visit, he was urgently hospitalized for aspiration pneumonia, which improved with antibiotic therapy. However, severe dysphagia persisted, making oral intake insufficient to maintain nutrition. Although he did not explicitly request tube feeding, a dysphagia diet was introduced. His respiratory function gradually declined despite medical intervention, and he died 45 days after admission three years after the first visit. An autopsy was conducted with the family’s consent. Representative PET and neuropathological findings have been reported previously [3].

### Neuroradiology

#### MRI studies

MRI was performed on a 3.0-T MAGNETOM Verio scanner (Siemens Healthcare). A T1-weighted gradient-echo sequence image (T1WI) was acquired for coregistration and segmentation of the PET images. The T1WIs were obtained in the sagittal plane with 1-mm slice thickness, echo time (TE) of 1.95 ms, repetition time (TR) of 2300 ms, inversion time (TI) of 900 ms, flip angle of 9.0°, field of view (FOV) of 250 mm, and matrix dimension of 512 × 512 × 176.

#### PET studies

The radiosynthesis of [^18^F]florzolotau was performed as previously described [9]. PET was conducted using a Biograph mCT Flow system (Siemens Healthcare) with a matrix dimension of 200 × 200 × 109 and a voxel size of 2 × 2 × 2 mm. Images were reconstructed with a filtered back-projection algorithm employing a Hanning filter with a full width at half maximum of 6.0 mm. The detailed PET acquisition protocol has been reported elsewhere [4]. The patient underwent dynamic PET scanning between 90–110 min (frames, 2 × 10 min) after an intravenous injection of [^18^F]florzolotau (188.6 MBq). The molar activity of the radioligand at the time of injection was 213.2 GBq/μmol.

#### Image data preprocessing

Image preprocessing was conducted using PMOD 4.3 (PMOD Technologies LLC, Switzerland), Statistical Parametric Mapping software (SPM12; Wellcome Department of Cognitive Neurology), and M-Vision Brain (M Corporation, Japan).

T1WI was downsampled from a matrix dimension of 512 × 512 × 176 to 256 × 256 × 176 using M-Vision Brain, and a segmentation map was generated. Subsequently, motion-corrected and averaged [^18^F]florzolotau PET images were converted to SUVR (standardized uptake value ratio) images. The reference region for SUVR quantification was automatically extracted from gray matter using an in-house script implemented in MATLAB script (The MathWorks, Natick, MA, USA) based on histogram analysis, as described elsewhere [3].

The downsampled T1WI, segmentation map, and SUVR images were then subjected to diffeomorphic normalization to the Montreal Neurological Institute (MNI) space. This normalization employed a skull-stripped T1 MR template, implemented in PMOD (derived from SPM5) and used a symmetric normalization (SyN) algorithm from Advanced Normalization Tools (ANTS) integrated within PMOD.

#### Definition of regions of interest

SUVR measurements were performed in regions of interest (ROIs) derived from multiple anatomical atlases, including Automated Anatomical Labeling Atlas 3 (version 1), the SPM Anatomy Toolbox (version 3.0), and the Wake Forest University Pick Atlas, all in the MNI space, as well as regions segmented by M-Vision Brain. Since the pontine and medullary nuclei are relatively small, ventral and dorsal portions containing these nuclei were manually combined into single ROIs (see Supplementary Figure 1 and Table for detailed ROI specifications).

All SUVRs were measured on the left hemisphere to correspond with the side examined in the neuropathological assessment of tau burden.

#### PET results

Parametric [^18^F]florzolotau SUVR maps are shown in Figure 2. Markedly increased tracer uptake was observed in the midbrain, thalamus, subthalamic nucleus, basal ganglia, and choroid plexus. Mild to moderate elevations were also noted in the pons, cerebellar white matter, dentate nucleus of the cerebellum, and frontal white matter.

**Figure 2.**
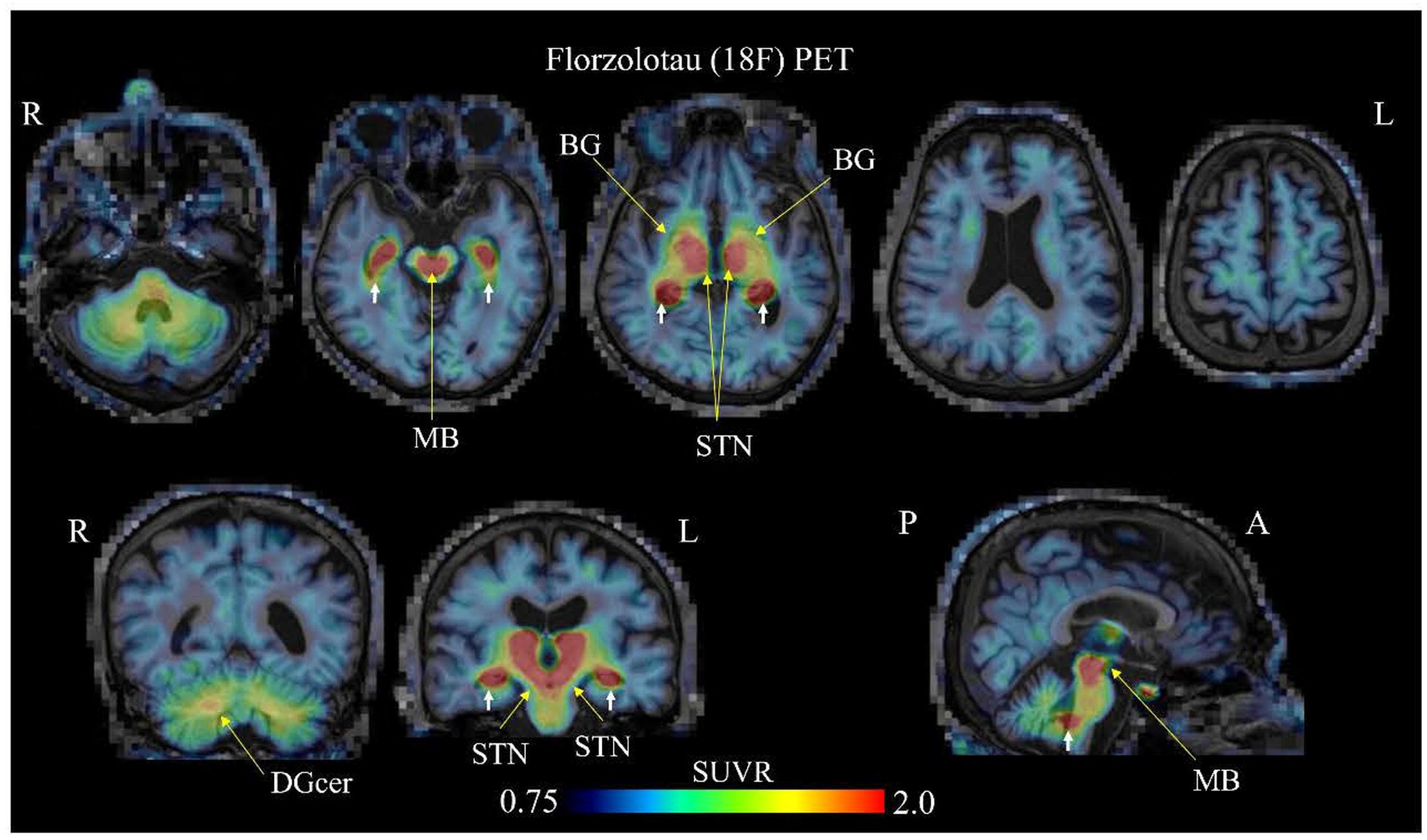
[^18^F]florzolotau PET findings. Parametric SUVR maps demonstrate pronounced tracer uptake in the upper pons-midbrain (MB), bilateral diencephalic structures including the subthalamic nucleus (STN), portions of the basal ganglia (BG), dentate nucleus of the cerebellum (DGcer), and choroid plexus (white arrow). Moderate uptake is observed in the middle pons to cerebellar white matter, whereas the frontal white matter shows mild uptake. SUVR, standardized uptake value ratio. Abbereviations: A: anterior, L: left, P: posterior, R: right.

### Neuropathology

#### Neuropathological examination

The unfixed brain weighed 1,200 g. The left hemisphere and spinal cord were fixed in formalin for four months. Gross examination revealed mild frontal lobe atrophy and marked atrophy of the midbrain tegmentum. Severe depigmentation was observed in the substantia nigra and locus coeruleus. The cerebellar cortex displayed mild atrophy with an indistinct outline of the dentate nucleus and prominent atrophy of the hilum (Figure 3).

**Figure 3.**
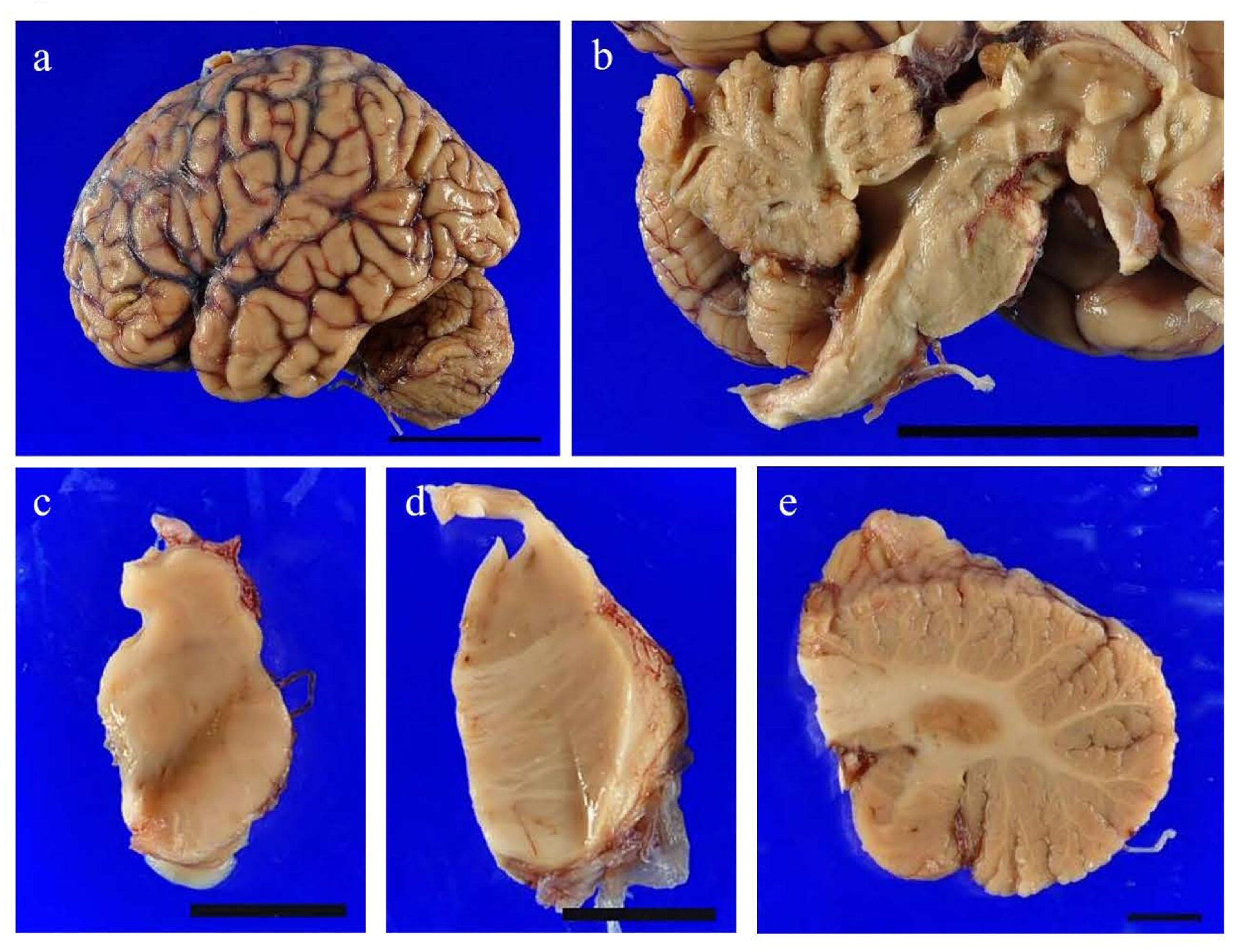
Macroscopic neuropathology. Gross examination reveals mild frontal lobe atrophy (a) and marked atrophy of the brainstem tegmentum with moderate dilation of the midbrain aqueduct (b). Pronounced depigmentation is observed in the substantia nigra (c) and locus coeruleus (d). The dentate nucleus appears indistinct, and the hilum is severely atrophic (e). Scale bars = 5 cm (a, b); 1 cm (c-e).

Tissues from 48 regions of the central nervous system were sampled, embedded in paraffin, and sectioned at 5 μm thickness. Sections were stained with hematoxylin-eosin, Luxol Fast Blue/Nissl (Klüver-Barrera), and Gallyas silver impregnation. Immunohistochemistry was performed using monoclonal antibodies against phosphorylated tau (AT-8; dilution, 1:500; CosmoBio, Tokyo, Japan) [10], α-synuclein (pSyn#64; dilution, 1:10,000; FUJIFILM Wako Pure Chemical Corp., Osaka, Japan) [11], and Aβ (BAN50; dilution, 1:2,000; FUJIFILM Wako Pure Chemical Corp., Osaka, Japan) [12], as well as a rabbit polyclonal antibody against phosphorylated TDP-43 (pS409/410; dilution, 1:3,000; CosmoBio, Tokyo, Japan) [13]. For α-synuclein and Aβ staining, formic acid pretreatment was applied for 5 min. Following incubation with a biotinylated secondary antibody (dilution, 1:500; Vector Laboratories, Burlingame, CA) and an avidin-biotinylated enzyme complex (Vectastain Elite ABC kit; Vector Laboratories, Burlingame, CA), signals were visualized with 3,3′-diaminobenzidine (DAB; 0.67 mg/mL; Sigma-Aldrich Inc., St. Louis, MO) and counterstained with hematoxylin.

#### Quantitative analysis of tau pathology

Quantification of tau burden was carried out on AT-8-immunostained sections. Five randomly selected microscopic areas were analyzed per region (three for the Meynert basal and subthalamic nuclei because of limited tissue availability) at 200x magnification under uniform conditions (BX53 microscope with DP74 camera; Olympus, Tokyo, Japan). AT-8-positive areas (DAB-stained) were determined by binary thresholding at a pixel intensity of 105/256. This threshold was established from histograms of 100 DAB-positive and 100 DAB-negative structures (Supplementary Figure 2). The percentage area of AT-8-positive pixels (%area) was calculated using Photoshop CC (Adobe Inc., San Jose, CA, USA) (Figure 4).

**Figure 4.**
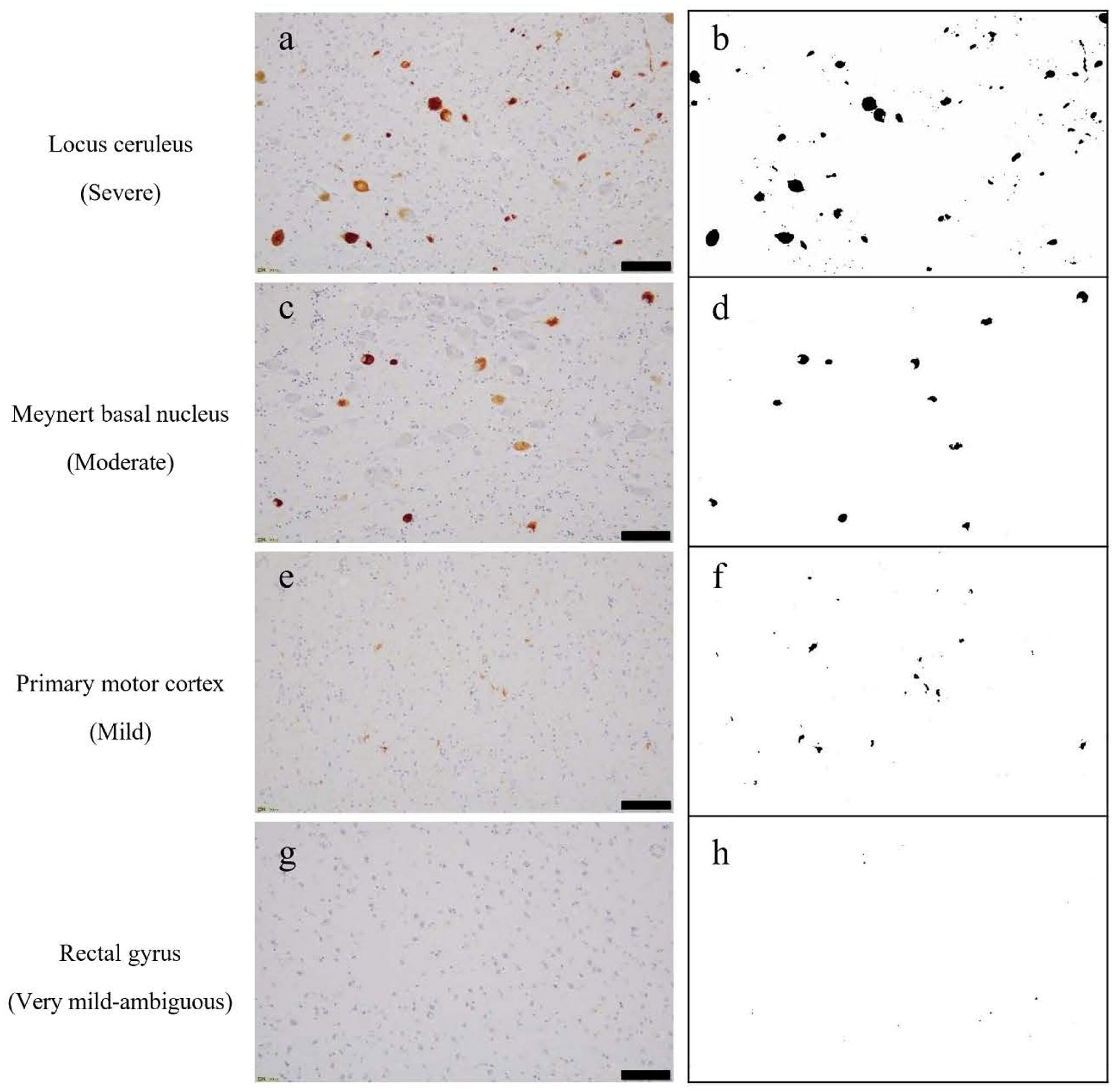
Quantification of tau burden. Microscopic images before (a, c, e, g) and after (b, d, f, h) binarization of AT-8 immunostaining are displayed. Tau burden was classified into four categories based on the percentage of AT-8-positive area: very mild/ambiguous (<0.1%), mild (0.1-0.2%), moderate (0.2-0.3%), and severe (≥0.3%). Scale bars = 100 μm.

Neuronal loss and gliosis were semi-quantitatively graded as follows: -, absent or negligible; +, mild; ++, moderate; and +++, severe.

#### Pathological findings

The regional severity of tau burden and neuronal loss/gliosis is summarized in Table 1, with representative mapping data displayed in Figure 5. AT-8- and Gallyas-positive neurofibrillary tangles (NFTs), tufted astrocytes, and neuropil threads were abundant in subcortical areas, including the locus coeruleus, substantia nigra, subthalamic nucleus, and midbrain-pontine tegmentum, whereas the cerebral cortices were relatively spared. The dentate nucleus exhibited marked neuronal loss accompanied by severe gliosis and microglial infiltration in the hilum. Tau deposition in the cerebellar cortex was minimal, and Purkinje cells were largely preserved. Numerous glial tau aggregates were present in the cerebellar white matter (Figure 6).

**Figure 5.**
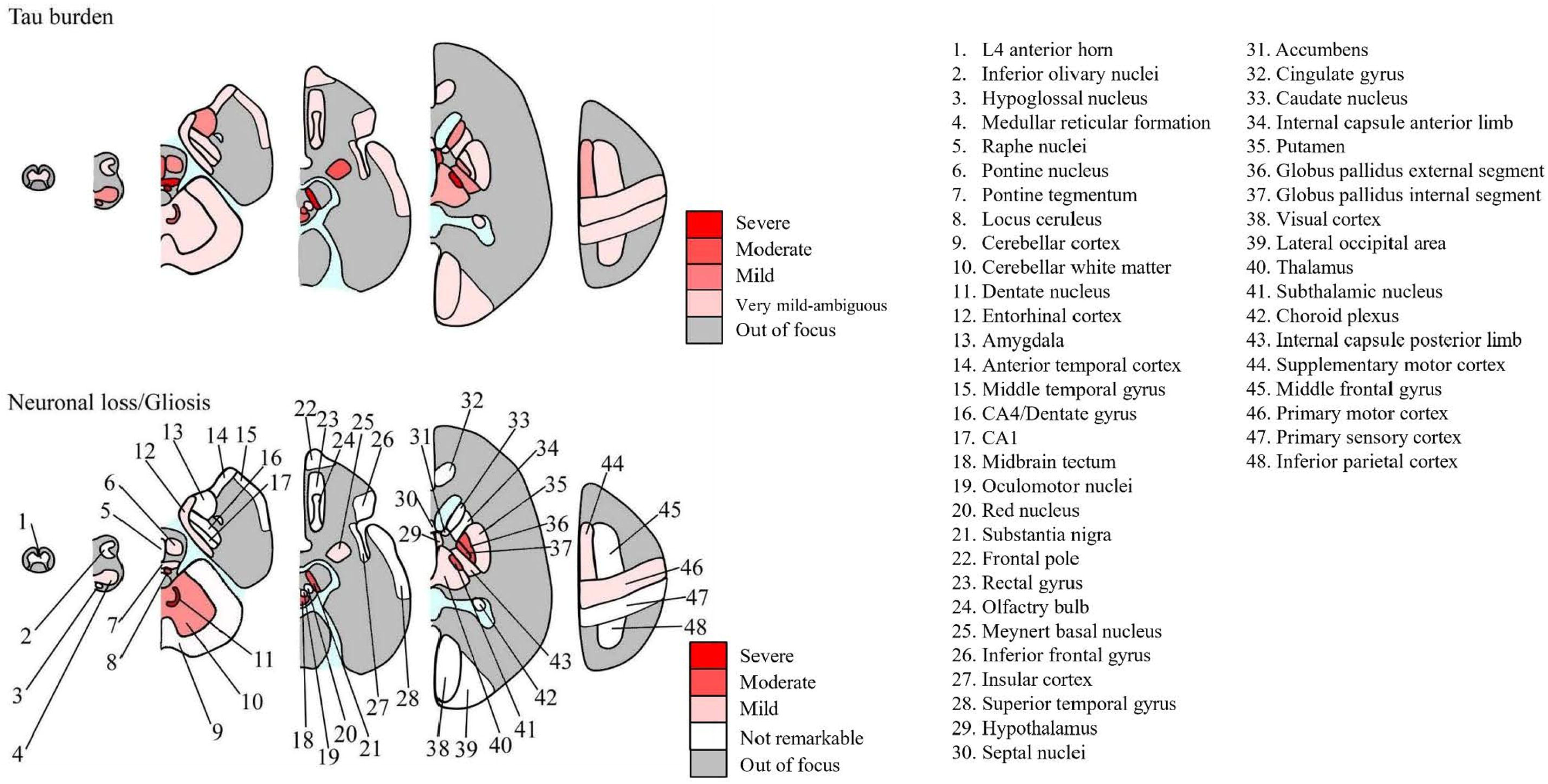
Schematic mapping of tau burden and neuronal loss/gliosis (NL/gliosis). Regional severity is color-coded using four red intensity grades. Tau burden severity follows the classification described in Figure 4. NL/gliosis was graded as unremarkable, mild, moderate, or severe.

**Figure 6.**
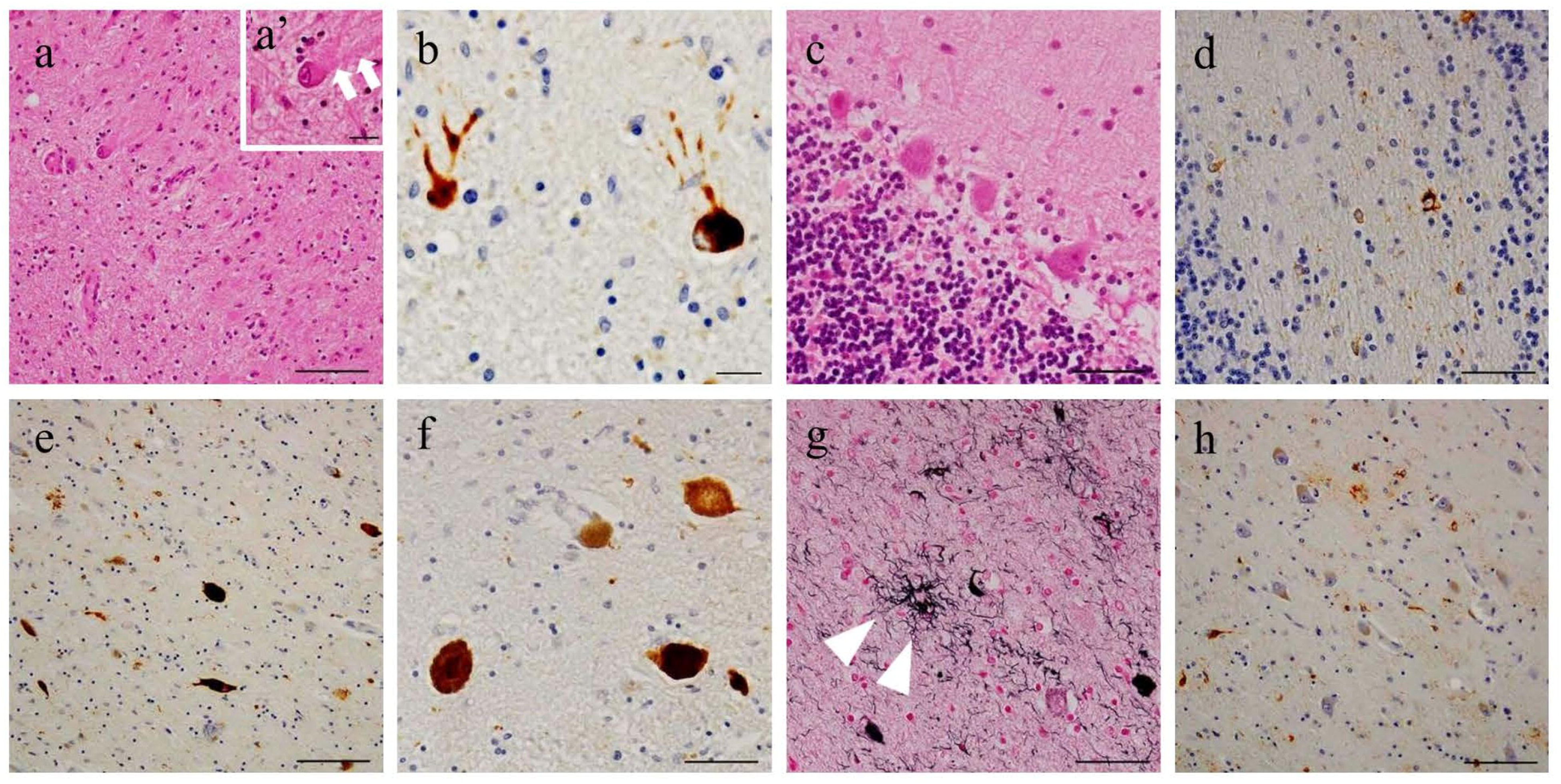
Microscopic tau pathologies. (a) Severe neuronal loss and gliosis in the dentate nucleus. (a’) Grumose degeneration around dendrites of remaining neurons (arrows). (b) Tau inclusions in remaining neurons. (c) Purkinje cells in the cerebellar cortex. (d) Abundant glial tau inclusions in the cerebellar white matter. (e, f) Tau-positive neuronal aggregates in the substantia nigra (e) and locus coeruleus (f). (g) Tufted astrocytes (arrowheads) and neuropil threads in the subthalamic nucleus (g). (h) Tau-positive aggregates in the thalamus (h). Staining: hematoxylin-eosin (a, c), AT-8 (b, d, e, f, h), Gallyas silver (g). Scale bars = 100 μm (a, e, h); 50 μm (c, d, f, g); 20 μm (a’, b).

**Table 1.**
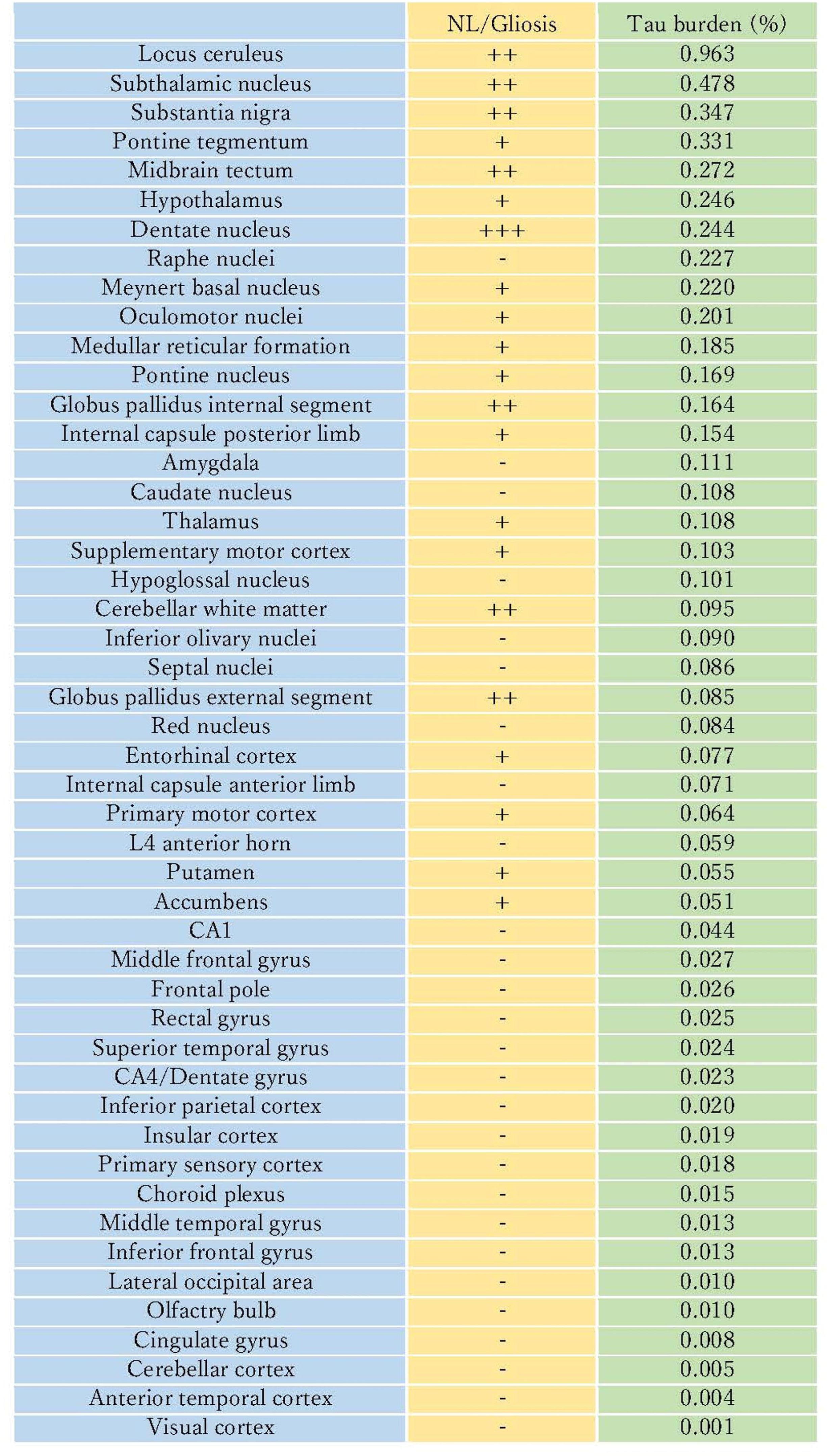
Quantitative analysis of tau burden and semiquantitative grading of neuronal loss and gliosis. NL, neuronal loss.

NFTs were detected in the entorhinal cortex and CA1 region (Braak NFT Stage II) [14]. Aβ plaques were confined to the neocortex and limbic areas (Thal phase 2) [15]. No Lewy pathology or TDP-43-positive inclusions were identified.

### Tau PET-neuropathology correlation analysis

The distribution of [^18^F]florzolotau uptake in the central nervous system closely mirrored the regional pattern of tau accumulation. The quantitative relationship between radioligand SUVRs and histopathological tau burden is shown in Figure 7. Statistical significance of this correlation was evaluated using Spearman’s rank correlation test, with p <0.05 considered significant.

**Figure 7.**
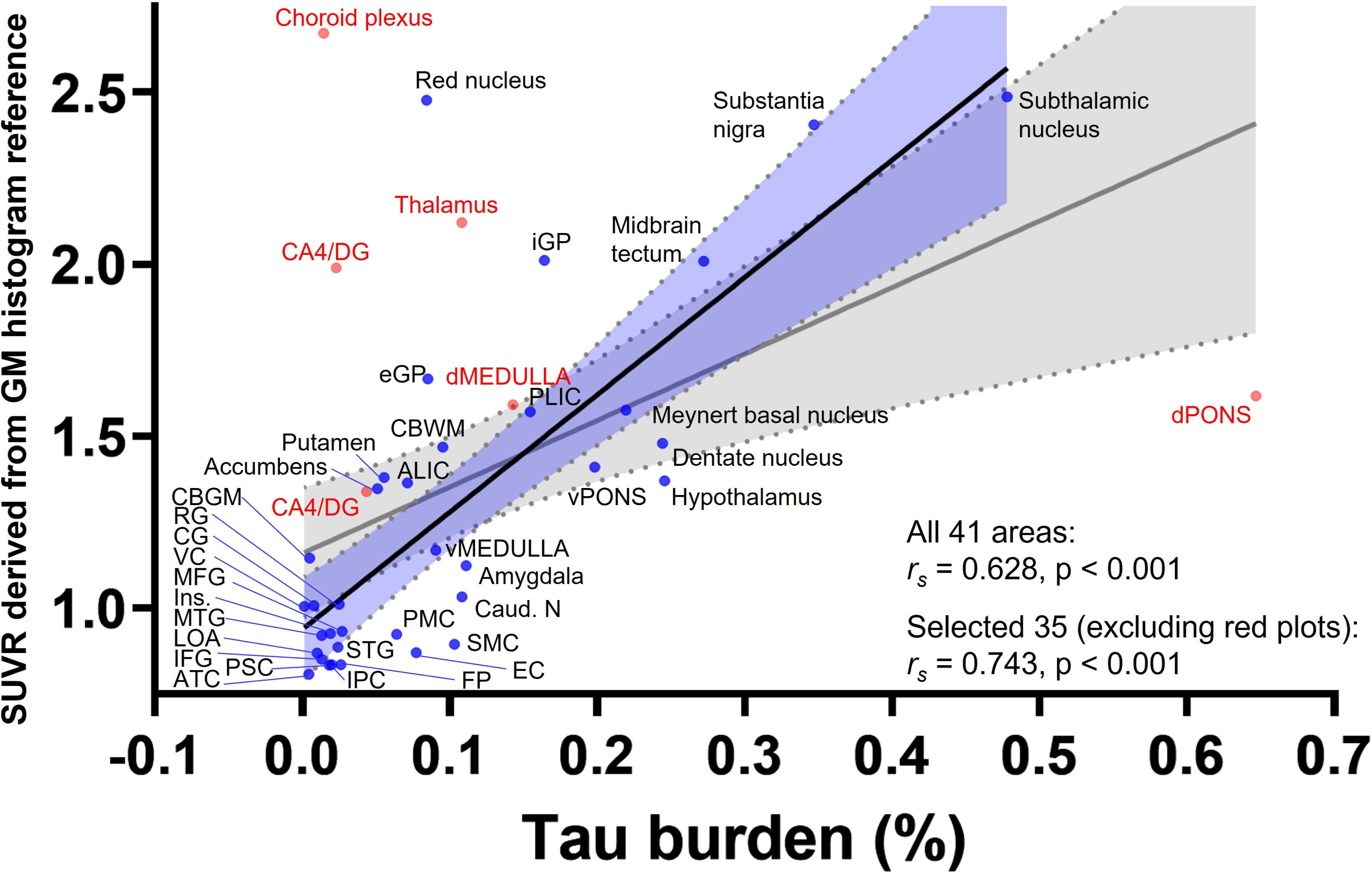
Correlation between [^18^F]florzolotau SUVR and histopathological tau burden. Scatter plots show the relationship between PET-derived SUVRs and tau burden across 41 regions (*r_s_* = 0.628, 95% confidence interval [CI]: 0.3874–0.7884, two-tailed p < 0.001). In this analysis, red plots indicate six regions with potential spill-in effects from the choroid plexus. After excluding these regions, the correlation increased (*r_s_* = 0.743, 95% CI: 0.5365–0.8650, p < 0.001; n = 35). For the overall correlation, the regression line is shown in gray with gray-shaded error bars representing the 95% CI. For the correlation excluding the red plots, the regression line is shown in black with light-blue shaded error bars representing the 95% CI. ALIC, internal capsule anterior limb; ATC, anterior temporal cortex; CA4/DG, CA4/Dentate gyrus; Caud. N, caudate nucleus; CBGM, cerebellar gray matter; CBWM, cerebellar white matter; CG, cingulate gyrus; dMEDULLA, dorsal medulla; dPONS, dorsal pons; EC, entorhinal cortex; eGP, globus pallidus external segment; FP, frontal pole; GM, gray matter; IFG, inferior frontal gyrus; iGP, globus pallidus internal segment; Ins., insular cortex; IPC, inferior parietal cortex; LOA, lateral occipital area; MFG, middle frontal gyrus; MTG, middle temporal gyrus; PLIC, internal capsule posterior limb; PMC, primary motor cortex; PSC, Primary sensory cortex; RG, rectal gyrus; SMC, supplementary motor cortex; STG, superior temporal gyrus; VC, visual cortex; vMEDULLA, ventral medulla; vPONS, ventral pons.

Correlation analyses were performed across 41 examined regions and, separately, across 35 regions after excluding six areas (including the dorsal medulla, dorsal pons, thalamus, and hippocampal subfields CA1 and CA4/dentate gyrus) where the radioactivity spill-in from the choroid plexus was evident. In both analyses, a strong and statistically significant association was observed between regional SUVR values and tau burden (41 regions: rs = 0.628, p < 0.001; 35 regions: rs = 0.743, p < 0.001).

Notably, increased radioligand uptake in the choroid plexus appeared unrelated to tau pathology. Histological examination revealed minimal AT-8-positive tau deposition in the this structure, suggesting that the elevated signal may reflect off-target binding [16].

## Discussion

This study represent the first successful demonstration of a topographical comparison between in vivo [^18^F]florzolotau PET findings and postmortem tau pathology in the PSP brain. A strong correlation was observed between the distribution of phosphorylated tau pathology revealed by histopathological analysis and the degree of [^18^F]florzolotau retention quantified by SUVR, underscoring the accuracy of PET with this tracer in capturing the spatial and quantitative characteristics of tau deposition.

Earlier generations of tau PET ligands faced several limitations, including low sensitivity for frontotemporal lobar degeneration (FTLD)-related tau lesions and non-specific white matter binding. [^18^F]florzolotau has largely overcome these challenges, demonstrating robust binding to both AD-type and non-AD tau pathologies such as PSP, CBD, and Pick’s disease. The tracer uptake have been shown to correlate with the clinical severity of AD and FTLD, and its performance distinguishes AD and PSP from healthy controls with sensitivity and specificity exceeding 90% [4].

In the present case, [^18^F]florzolotau PET findings closely mirrored the postmortem distribution and regional burden of tau pathology, confirming the reliability of this tracer for evaluating lesion spread and severity in PSP. These results provide compelling in vivo-to-postmortem validation of [^18^F]florzolotau as a sensitive and specific imaging biomarker for 4-repeat tauopathies.

Clinically, the diagnosis of PSP-C is often challenging due to overlapping features with other disorders. This case highlights the diagnostic potential of [^18^F]florzolotau PET in differentiating PSP from other parkinsonian or ataxic syndromes. Although the initial presentation of the patient with prominent limb ataxia did not fully meet the MDS-PSP clinical criteria, the coexistence of vertical supranuclear gaze palsy and frequent backward falls within three years of onset supported a diagnosis of PSP [8]. Despite minimal cerebellar cortical tau pathology or Purkinje cell loss, it is plausible that early ataxic symptoms reflected dysfunction of the cerebellar dentate nucleus and its efferent pathways, followed by progressive involvement of the midbrain and globus pallidus. Over time, parkinsonian features such as rigidity masked the ataxic manifestations.

Several limitations should be acknowledged. First, as it is a single case study, further clinicopathological investigations are required to confirm the generalizability of these findings. Second, off-target retention of [^18^F]florzolotau to the choroid plexus, presumably arising from interactions of this tracer with Tmem106B aggregates within this structure [17], may affect quantitative accuracy in adjacent regions such as the medulla tegmentum, hippocampus, and dorsal thalamus.

In conclusion, [^18^F]florzolotau PET provides a powerful means to visualize and quantify 4-repeat tau deposition in vivo. Along with previous evidence in Pick’s disease [5], this neuroimaging technology allows detailed assessments of diverse primary tauopathies. Its high topographical accuracy and pathological validity highlight its value for clinical diagnosis, trial stratification, and therapeutic evaluation in patients with PSP and other FTLD-tau disorders.

## Data Availability

All data produced in the present work are contained in the manuscript.

## Availability of data and materials

The study’s clinical, radiological, and neuropathological datasets are available from the Corresponding Author upon request.

## Abbreviations

PET: positron emission tomography PSP: progressive supranuclear palsy
PSP-C: PSP with predominant cerebellar ataxia CBD: corticobasal degeneration
SBR: specific binding ratio
SUVR: standardized uptake value ratio RI: radioisotope
TDP-43: transactive response DNA-binding protein 43kDa FTLD: frontotemporal lobar degeneration
AD: Alzheimer’s disease

## Acknowledgments

We extend our heartfelt gratitude to the patient and his family for their cooperation. We would like to thank the neurology members of the Department of Neurology, National Hospital Organization, Chiba Medical Center Chibahigashi National Hospital, and the Department of Neurology, Graduate School of Medicine, Chiba University, for their invaluable assistance with the clinical examination. Additionally, we are grateful to the radiology team at the Institute for Quantum Medical Science in the National Institutes for Quantum Science and Technology for their contributions to obtaining PET imaging. The exceptional pathological analyses conducted by the Department of Pathology, National Hospital Organization, Chiba Medical Center Chibahigashi National Hospital, and the Laboratory of Neuropathology, Tokyo Metropolitan Institute of Medical Science, are sincerely acknowledged.

## Authors’ information

Authors and affiliation

Department of Neurology, National Hospital Organization Chiba Medical Center Chibahigashi National Hospital, Chiba

673 Nitona-cho, Chuo-ku, Chiba, 260-8712 Japan

Nobuyuki Araki, Takahiro Takeda, Sagiri Isose, Kimiko Ito, Kimihito Arai and Kazuhiro Honda

Department of Neurology, Graduate School of Medicine, Chiba University, Chiba 1-8-1 Inohana, Chuo-ku, Chiba, 260-8670 Japan

Nobuyuki Araki and Satoshi Kuwabara

Institute for Quantum Medical Science, National Institutes for Quantum Science and Technology, Chiba

4-9-1 Anagawa, Inage-ku, Chiba, 263-8555

Hironobu Endo, Yuhei Takado, Kenji Tagai and Makoto Higuchi

Department of Psychiatry, The Jikei University of Medicine, Tokyo 105-8461, Japan 3-25-8 Nishishinbashi, Minato-ku, Tokyo, 105-8461, Japan

Kenji Tagai

Laboratory of Neuropathology, Tokyo Metropolitan Institute of Medical Science, Tokyo, Japan

2-1-6 Kamikitazawa, Setagaya-ku, Tokyo, 156-8506 Japan Erika Seki and Nobutaka Arai

Neuroetiology and Diagnostic Science, Osaka Metropolitan University Graduate School of Medicine, Osaka, Japan

1-4-3 Asahi-machi, Abeno-ku, Osaka 545-0051, Japan Makoto Higuchi

## Contributions

NA: Designed the research project, obtained the clinical information, analyzed the clinical data, and drafted the manuscript.

TT: Designed the research project, analyzed the pathological data, and drafted the manuscript.

YT: Designed the research project and obtained the radiological data. KT: Obtained the radiological data.

ES: Performed the pathological examination. NA: Performed the pathological examination. SI: Obtained the clinical information.

KI: Obtained the clinical information. KA: Interpreted the results.

KH: Obtained the clinical information and interpreted the results. SK: Supervised the study and revised the manuscript

MH: Supervised the study and revised the manuscript

HE: Designed the research project, obtained the radiological data, and drafted the manuscript.

## Corresponding authors

Correspondence is to be addressed to Nobuyuki Araki and/or Hironobu Endo

## Ethics declaration

### Ethics approval

All procedures performed in this report were following the ethical standards of the institutional research committees and the 1964 Helsinki Declaration and its later amendments. Written informed consent was obtained.

## Consent for publication

The patient’s family has consented to publication.

## Supplementary Table

List of regions of interest used for pathological assessment, corresponding PET regions, and atlas-based anatomical references.

**Supplementary Figure 1**

Anatomical illustration of ROIs corresponding to PET regions and atlas-based references. Abbreviations used in this figure are defined in the Supplementary Table.

**Supplementary Figure 2**

Histogram-based determination of the brightness threshold separating AT-8-positive from AT-8-negative structures for binary image analysis.

