## Supplementary figures and images for "Florzolotau (18F) PET provides in-vivo measures of tau pathology in progressive supranuclear palsy: An imaging-postmortem correlation study"

### Supplementary figure 1

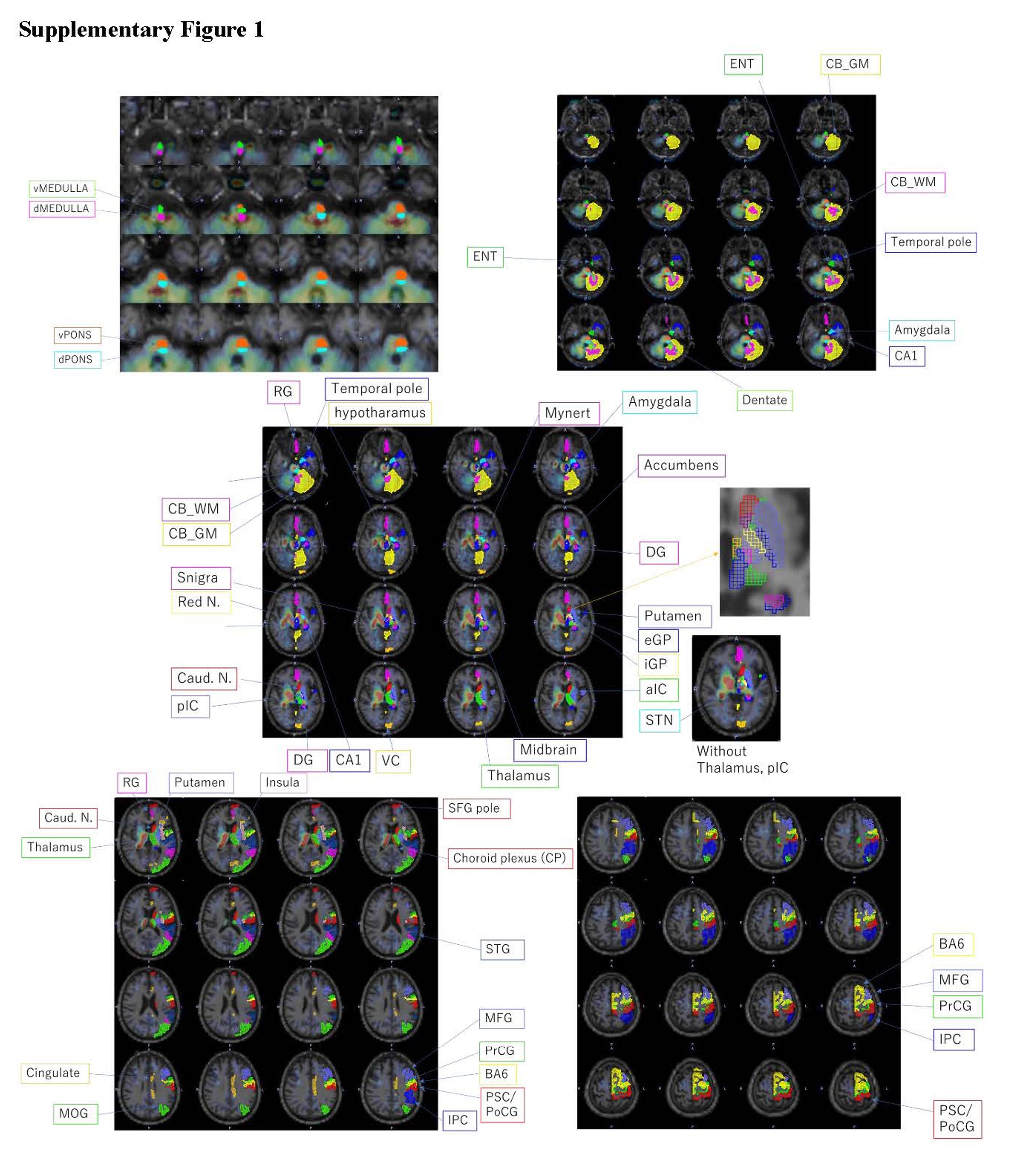

### Supplementary figure 2

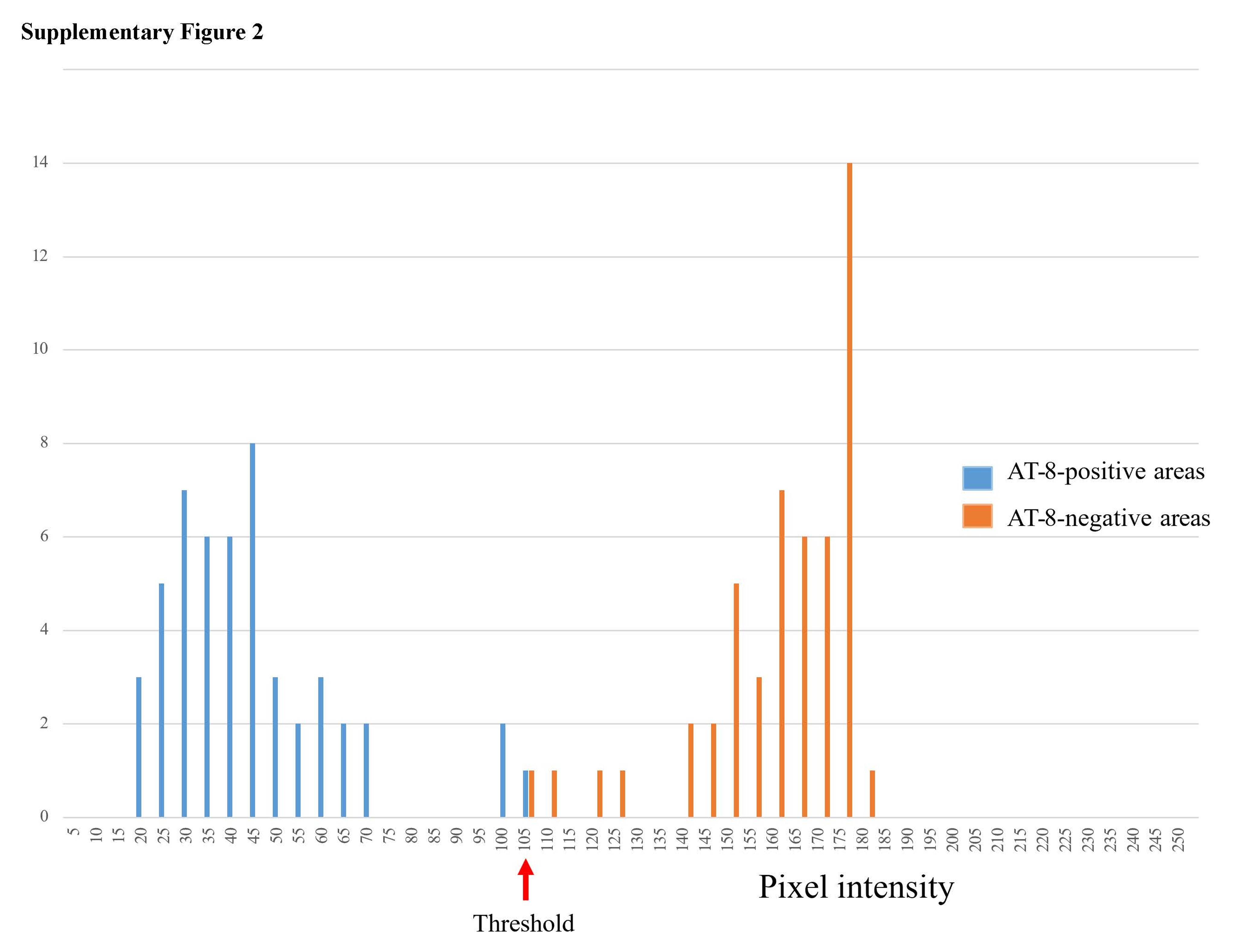

### Supplementary table

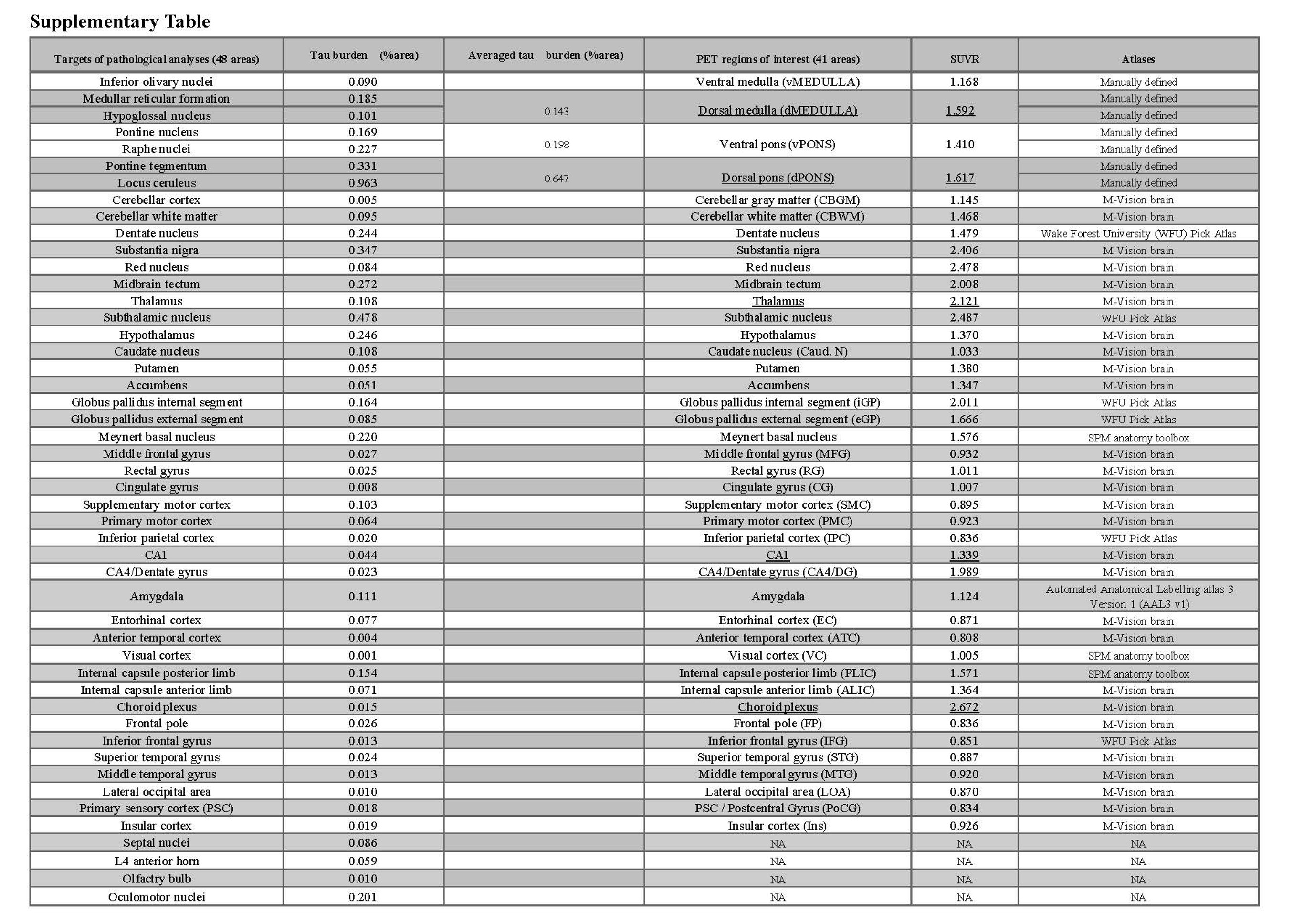
